# Spatial transcriptomic patterns underlying regional vulnerability to amyloid-β and tau pathologies and their relationships to cognitive dysfunction in Alzheimer’s disease

**DOI:** 10.1101/2023.08.12.23294017

**Authors:** Meichen Yu, Shannon L. Risacher, Kwangsik T. Nho, Qiuting Wen, Adrian L. Oblak, Frederick W. Unverzagt, Liana G. Apostolova, Martin R. Farlow, Jared R. Brosch, David G. Clark, Sophia Wang, Rachael Deardorff, Yu-Chien Wu, Sujuan Gao, Olaf Sporns, Andrew J. Saykin, the Alzheimer’s Disease Neuroimaging Initiative (ADNI)

## Abstract

Amyloid-β (Aβ) and tau proteins accumulate within distinct neuronal systems in Alzheimer’s disease (AD). Although it is not clear why certain brain regions are more vulnerable to Aβ and tau pathologies than others, gene expression may play a role. We studied the association between brain-wide gene expression profiles and regional vulnerability to Aβ (gene-to-Aβ associations) and tau (gene-to-tau associations) pathologies leveraging two large independent cohorts (n = 715) of participants along the AD continuum. We identified several AD susceptibility genes and gene modules in a gene co-expression network with expression profiles related to regional vulnerability to Aβ and tau pathologies in AD. In particular, we found that the positive *APOE*-to-tau association was only seen in the AD cohort, whereas patients with AD and frontotemporal dementia shared similar positive *MAPT*-to-tau association. Some AD candidate genes showed sex-dependent negative gene-to-Aβ and gene-to-tau associations. In addition, we identified distinct biochemical pathways associated with the gene-to-Aβ and the gene-to-tau associations. Finally, we proposed a novel analytic framework, linking the identified gene-to-pathology associations to cognitive dysfunction in AD at the individual level, suggesting potential clinical implication of the gene-to-pathology associations. Taken together, our study identified distinct gene expression profiles and biochemical pathways that may explain the discordance between regional Aβ and tau pathologies, and filled the gap between gene-to-pathology associations and cognitive dysfunction in individual AD patients that may ultimately help identify novel personalized pathogenetic biomarkers and therapeutic targets.

**One Sentence Summary:** We identified replicable cognition-related associations between regional gene expression profiles and selectively regional vulnerability to amyloid-β and tau pathologies in AD.

## INTRODUCTION

Abnormal accumulations of amyloid-β (Aβ) and tau proteins in the brain are two principal neuropathological hallmarks of Alzheimer’s disease (AD). Aβ and tau pathologies affect distinct neuronal systems, leading to neurodegeneration and cognitive decline in AD^1–3^. Using positron emission tomography (PET), many studies have reported that Aβ initially accumulates in the medial frontal cortex and medial parietal cortex^4^, both parts of the default mode network^5,6^. In contrast, tau is initially deposited in the medial temporal lobe memory system, spreading from the transentorhinal cortex to the hippocampus and parahippocampal cortex, and finally to other brain regions^7–10^. However, why distinct brain regions are more selectively vulnerable to Aβ and tau pathologies than others remain to be elucidated.

The recent development of brain-wide gene expression atlas, e.g., the Allen Human Brain Atlas (AHBA^11,12^), has made it possible to connect spatial variations in gene expression profiles to the regional vulnerability to Aβ and tau pathologies (gene-to-pathology associations) in AD^13,14^. For instance, three recent studies^15–17^ have identified genes whose expression profiles are related to the spatial accumulation patterns of Aβ and tau pathologies. Additionally, in these studies, Aβ and tau-related biochemical pathways have also been reported. However, results from these studies are not always consistent, possibly due to the use of different approaches for processing the gene expression data and the use of PET imaging data with small sample size (n < 100). Recent large-scale GWAS studies^18–20^ have discovered many novel AD risk loci providing clues to molecular mechanisms, yet the potential of these risk loci to inform gene-to-pathology associations has not yet been studied. Moreover, Aβ pathology is specific to AD, whereas tau pathology is shared in different types of dementia, for example, frontotemporal lobar degeneration (FTLD). It is not clear if the regional vulnerability to tau pathology and its relation to regional gene expression profiles were specific to AD or shared with FTLD. Finally, until now, the gene-to-pathology associations remain uncertain at the individual level and have not been related to cognitive dysfunction in AD patients, hindering their translation to clinical utility.

To fill these gaps, we tested the hypotheses that brain-wide gene expression profiles are associated with selective regional vulnerability to Aβ and tau pathologies, and that these spatial gene-to-pathology associations are related to cognitive dysfunction in AD. We used two large independent datasets, namely the Alzheimer’s Disease Neuroimaging Initiative (ADNI; n = 605) and Indiana Memory and Aging Study (IMAS; n = 110) cohorts. Specifically, we used the ADNI cohort as a discovery dataset, and then used the IMAS dataset to replicate the ADNI findings. Regional gene expression profiles for 15,745 protein-coding genes were derived from brain-wide microarray-based transcriptome data from the AHBA. In this report, we conducted one hypothesis-driven analysis using a priori selected AD susceptibility genes selected from recent large-scale GWAS studies^18–20^ and two data-driven analyses using all 15,745 genes. A schematic overview of the applied methods is provided in **Fig. 1**. First, in the hypothesis-driven analysis (**Fig. 1A**), we studied how gene expression profiles of individual AD susceptibility genes are related to regional vulnerability to Aβ (gene-to-Aβ associations) and tau (gene-to-tau associations) pathologies. We tested if the gene-to-tau associations are specific to AD or shared with FTLD by re-conducting the gene-to-tau association analysis in 11 patients with FTLD. Spatial permutation testing and gene specificity testing were performed to verify the robustness of the identified gene-to-pathology association. In a subsequent data-driven analysis (**Fig. 1B**), we computed spatial gene-to-amyloid-β and gene-to-tau associations for average gene expression profiles of gene modules identified from a gene co-expression network (15,745 × 15,745). Then, in another data-driven analysis (**Fig. 1C**), we identified biochemical pathways that may underlie the regional vulnerability to Aβ and tau pathologies using gene set enrichment analysis (GSEA^21^) of the full genome-wide expression profiles. Finally, we developed a novel analytic framework for estimating how cognitive impairments in AD are related to the identified gene-to-Aβ and gene-to-tau associations at the levels of individual genes and gene co-expression modules (**Fig. 1D**).

**Figure 1.**
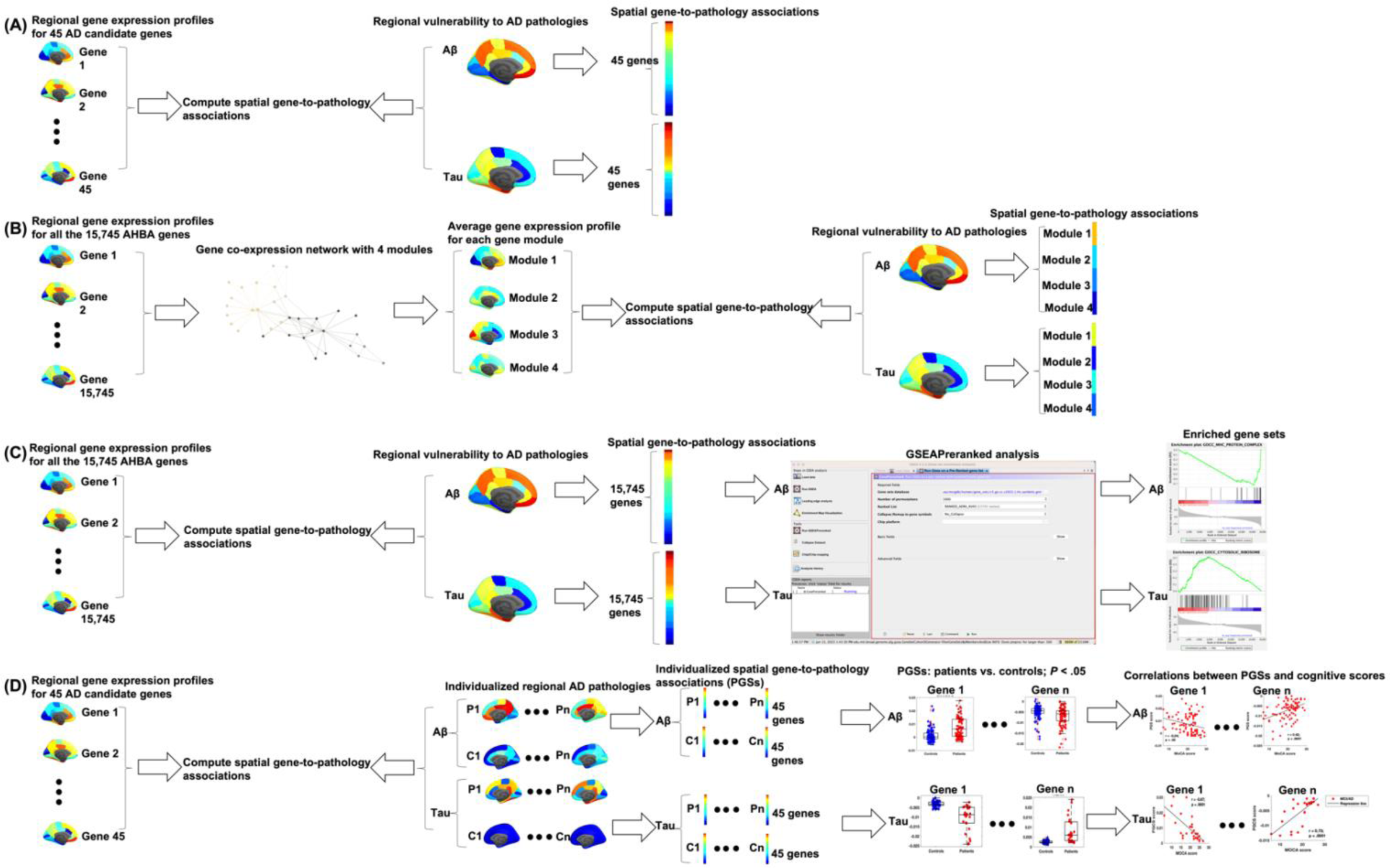
Schematic overview of the applied methods. **(A)** In a hypothesis-driven analysis, we computed spatial gene-to-pathology associations between brain-wide gene expression profiles of 45 AD susceptibility genes and regional vulnerability to Aβ as well as tau pathologies. We then performed two data-driven analyses **(B, C)** using regional gene expression profiles of all the 15,745 AHBA genes. **(B)** We constructed a gene co-expression network, where nodes represent genes and links denote correlations between expression profiles of 15,745 genes. We then identified 4 gene modules and compute spatial associations between average gene expression profiles of the 4 gene modules and regional vulnerability to Aβ as well as tau pathologies. **(C)** We computed spatial associations between brain-wide gene expression profiles of 15,745 genes and regional vulnerability to Aβ as well as tau pathologies. We then used the GSEA software to conduct GSEAPreranked analysis on the ranked spatial correlations and identified enriched gene sets. **(D)** We computed personalized spatial gene-to-pathology associations between brain-wide gene expression profiles of 45 AD susceptibility genes and individualized regional Aβ and tau pathologies, resulting in individual-level Aβ- and tau-related pathogenetic scores (PGSs). Then, we estimated group differences of PGSs between patients (MCI and AD) and controls (CU and/or SCD) for each gene. Finally, for the genes showing significantly different PGSs between groups, we computed Pearson’s correlations between the PGSs and cognitive scores (e.g., MoCA and MMSE). The above analyses **(A-D)** were performed using both the ADNI (discovery) and IMAS (replication) datasets. Analysis **(D)** was also performed using average expression profiles of 4 gene modules identified in analysis (B). Abbreviations: CU = cognitively unimpaired; SCD = subjective cognitive decline; MCI = mild cognitive impairment; AD = Alzheimer’s disease; IMAS = Indiana Memory and Aging Study; ADNI = Alzheimer’s Disease Neuroimaging Initiative; MoCA = Montreal Cognitive Assessment; MMSE = Mini-Mental State Exam; GSEA = gene set enrichment analysis.

## RESULTS

### Demographics

Participant characteristics and cognitive scores are presented in **Table 1**. The ADNI sample includes 336 cognitively unimpaired (CU) individuals, 200 patients with mild cognitive impairment (MCI), and 69 patients with AD dementia. The IMAS cohort includes 38 CU, 30 MCI, and 11 AD individuals, as well as 31 individuals with subjective cognitive decline (SCD). For the ADNI cohort, age and sex showed significant group differences (*P* < .0001); for the IMAS cohort, they did not differ across the four groups. Cognitive function of participants was assessed by both the Montreal Cognitive Assessment (MoCA) and Mini-Mental State Exam (MMSE) for the ADNI cohort and the MoCA for the IMAS cohort. The mean MoCA and MMSE scores were significantly (*P* < .0001) lower in patients (MCI and AD) compared to the psychometrically unimpaired groups (CU and/or SCD) in both cohorts.

**Table 1.** Characteristics of ADNI and IMAS samples.

|  | ADNI |  |  |  | IMAS |  |  |  |  |
| --- | --- | --- | --- | --- | --- | --- | --- | --- | --- |
|  | CU | MCI | AD | *P* | CU | SCD | MCI | AD | *P* |
| n | 336 | 200 | 69 | n.a. | 38 | 31 | 30 | 11 | n.a. |
| Age | 70.1 ±<br>6.81 | 71.7 ±<br>7.43 | 74.2 ±<br>8.50 | < .001 | 71.1 ±<br>6.19 | 70.9 ±<br>6.96 | 73.8 ±<br>7.11 | 71.9 ±<br>7.34 | .35 |
| Sex (F/M) | 198/138 | 94/106 | 322/283 | < .001 | 29/9 | 17/14 | 15/15 | 7/4 | .12 |
| MoCA | 26.0 ±<br>2.72 | 22.8 ±<br>3.40 | 16.5 ±<br>4.41 | < .001 | 26.4 ±<br>2.03 | 25.9 ±<br>2.53 | 20.5 ±<br>4.02 | 13.7 ±<br>4.61 | < .001 |
| MMSE | 29.1 ±<br>1.05 | 27.7 ±<br>2.05 | 22.8 ±<br>2.77 | < .001 | n.a. | n.a. | n.a. | n.a. | n.a. |

n = sample size; F = female; M = male; MoCA = Montreal Cognitive Assessment; MMSE = Mini-Mental State Exam; CU = cognitively unimpaired; SCD = subjective cognitive decline; MCI = mild cognitive impairment; AD = Alzheimer's disease; *P* = *P* values; n.a. = not available.
One-way ANOVA tests (for age, MoCA and MMSE) or chi-square ( $\chi^2$ ) tests (for sex) were used when applicable. Age, MoCA and MMSE scores are presented as mean and standard deviation.

### Regional vulnerability of Aβ and tau pathologies in AD

In both cohorts, Aβ pathology was measured using [^18^F]florbetapir (AV45) and [^18^F]florbetaben (FBB) PET; tau pathology was measured using [^18^F]flortaucipir (AV-1451) PET. We estimated the regional vulnerability to Aβ and tau pathologies by contrasting amyloid-β and tau PET imaging data between cognitively normal participants (CU in the ADNI cohort; CU and SCD in the IMAS cohort) and cognitively impaired patients (MCI and AD in two cohorts). In general, the ADNI and IMAS cohorts shared similar regional vulnerability to Aβ and tau pathologies: for Aβ, ADNI AV45 vs. ADNI FBB (Pearson’s rho = 0.84, *P* < .0001), ADNI AV45 vs. IMAS (Pearson’s rho = 0.65, *P* < .0001), ADNI FBB vs. IMAS (Pearson’s rho = 0.80, *P* < .0001); for tau, ADNI vs. IMAS (Pearson’s rho = 0.88, *P* < .0001). Specifically, in both cohorts, AD and MCI patients showed higher amyloid-β loads than CU and/or SCD, particularly in the medial parietal cortex, temporal lobe, medial and inferior prefrontal cortices, and superior and middle frontal cortices (**Fig. 2A**). The medial temporal lobe and visual cortex showed relatively lower Aβ levels than other brain regions in MCI and AD. In both cohorts, AD and MCI patients also showed higher tau levels than CU and/or SCD, mainly in the medial temporal lobe, medial and inferior parietal cortices, and inferior and middle temporal cortices (**Fig. 3A**). The frontal lobe and sensorimotor cortex showed relatively lower tau levels than other brain regions in MCI and AD. Females showed overall higher vulnerability to Aβ (**Fig. S2A**) and tau (**Fig. S3A**) pathologies across the cortex than males, although the spatial patterns of vulnerability were generally consistent in both sexes.

**Figure 2.**
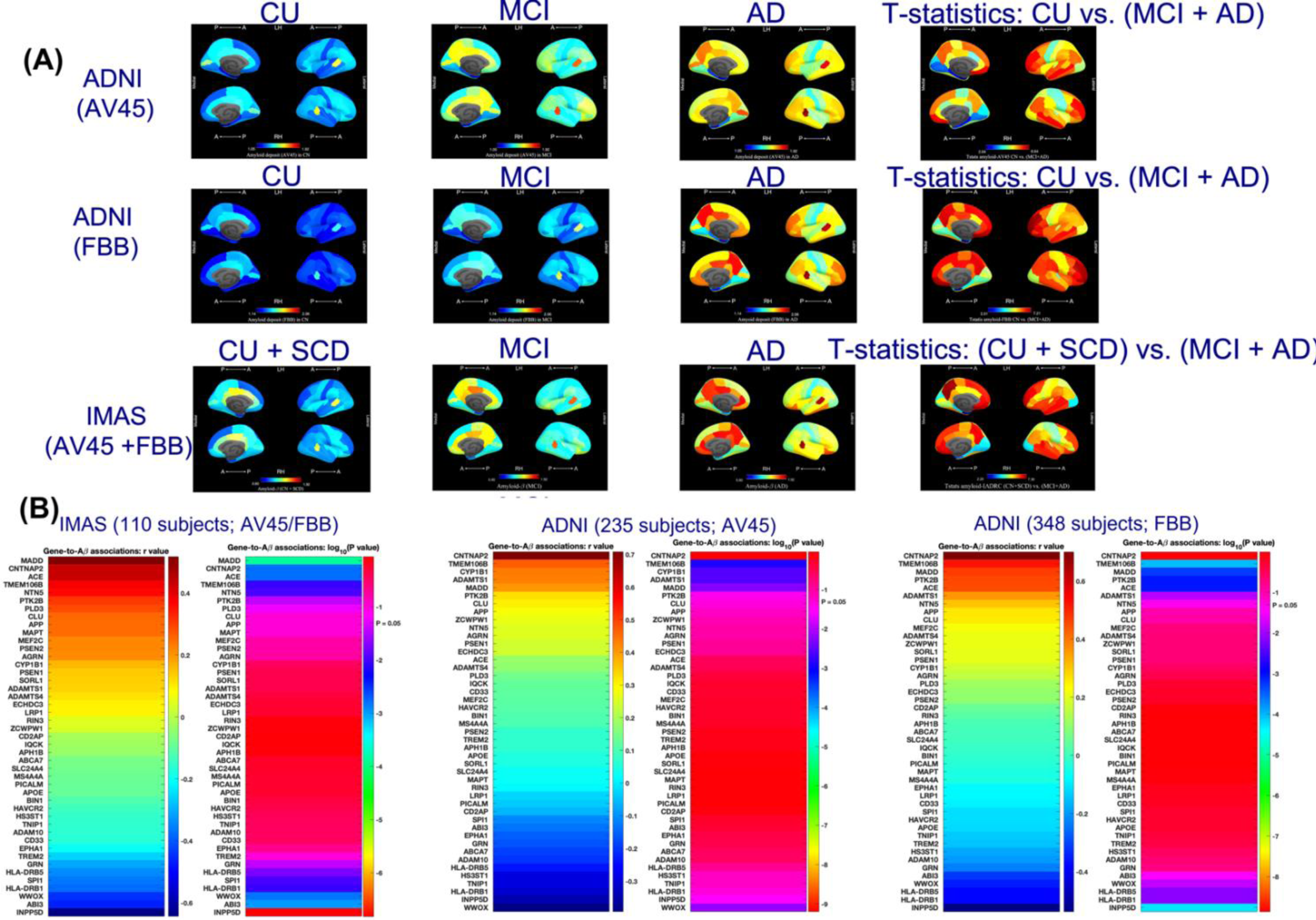
**(A)** Spatial patterns for Aβ deposits (SUVR values) across diagnosis groups and T-statistics map for regional differences. **(B)** Spatial gene-to-Aβ associations between brain-wide gene expression profiles of 45 AD susceptibility genes selected from recent large-scale GWAS studies and brain-wide Aβ data measured by two PET tracers in the IMAS and ADNI cohorts, separately. *P*-values were log10 transformed in **(B)**. Abbreviations: A = anterior; P = posterior; RH = right hemisphere; LH = left hemisphere; CU = cognitively unimpaired; SCD = subjective cognitive decline; MCI = mild cognitive impairment; AD = Alzheimer’s disease; IMAS = Indiana Memory and Aging Study; ADNI = Alzheimer’s Disease Neuroimaging Initiative; AV45 = [18F]florbetapir; FBB = [18F]florbetaben.

**Figure 3.**
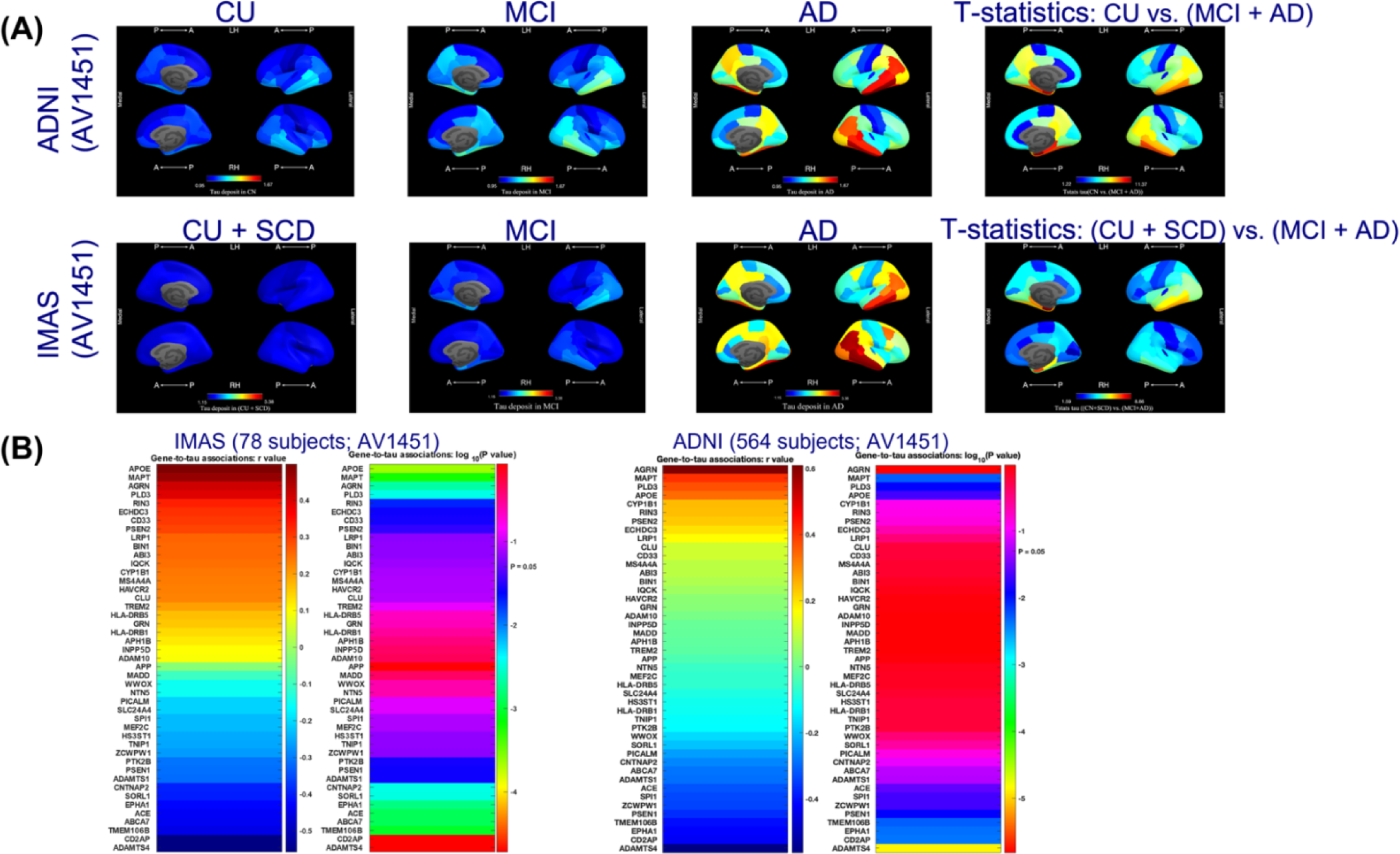
**(A)** Spatial patterns for tau deposits (SUVR values) across diagnosis groups and *T*-statistics map for regional differences. **(B)** Spatial gene-to-tau associations between brain-wide gene expression profiles of 60 AD susceptibility genes selected from recent large-scale GWAS studies and brain-wide tau PET data in the IMAS and ADNI cohorts, separately. *P*-values were log10 transformed in **(B)**. Abbreviations: A = anterior; P = posterior; RH = right hemisphere; LH = left hemisphere; CU = cognitively unimpaired; SCD = subjective cognitive decline; MCI = mild cognitive impairment; AD = Alzheimer’s disease; IMAS = Indiana Memory and Aging Study; ADNI = Alzheimer’s Disease Neuroimaging Initiative; AV1451 = [^18^F]flortaucipir.

### Genetic associations of Aβ and tau pathologies

In a hypothesis-driven candidate gene analysis, we computed spatial associations between gene expression profiles of 45 AD susceptibility genes and regional vulnerability to deposition of Aβ (gene-to-Aβ associations) and tau (gene-to-tau associations) pathologies (case-control *T*-statistic maps). In both cohorts, we identified consistent significant gene-to-Aβ and gene-to-tau associations after multiple testing adjustment for multiple genes (P_FDR_ < 0.05; (**Fig. 2B**; **Fig. 3B**)). Specifically, *CNTNAP2* and *TMEM106B* showed the strongest positive gene-to-Aβ associations, whereas *INPP5D*, *WWOX,* and *HLA-DRB1* showed the strongest negative associations in AD. In contrast, *APOE*, *MAPT, AGRN,* and *PLD3* showed the strongest positive gene-to-tau associations, whereas *ADAMTS4* and *CD2AP* showed the strongest negative associations.

Although Aβ pathology is specific to AD, other types of dementia can feature tauopathy, including some forms of frontotemporal lobar degeneration (FTLD). Therefore, we next tested if the gene-to-tau associations were specific to an AD population or were observed in patients with FTLD. We examined the pathological specificity of the identified gene-to-tau associations in 11 patients with FTLD from the IADRC cohort and found that the positive *APOE*-to-tau association and the negative *CD2AP*-to-tau association were only seen in the AD cohort, whereas AD and FTLD shared similar positive (e.g., *MAPT, AGRN,* and *PLD3*) and negative (e.g., *ADAMTS4*) gene-to-tau associations in the same genes (**Fig. S1**).

The identified positive gene-to-Aβ and gene-to-tau associations were generally consistent in males (**Fig. S2B**) and females (**Fig. S3B**). Some genes showed sex-dependent negative gene-to-Aβ and gene-to-tau associations. For instance, the *HLA-DRB1-* and *HLA-DRB5*-to-Aβ associations were more pronounced in males than in females, whereas the *WWOX*-to-Aβ association was only found in females. Similarly, the *CD2AP*-to-tau association was specific to females. The identified gene-to-Aβ and gene-to-tau associations were corrected for the effects of spatial autocorrelation and gene specificity (see **Supplement Table S1**). The gene-to-Aβ associations showed higher gene specificity than the gene-to-tau associations, which is consistent with our hypothesis and the results of pathological specificity analysis described above.

Going beyond candidate gene analysis using preselected individual genes to a data-driven analysis, we constructed a gene co-expression network by computing the Pearson correlations of gene expression profiles between all pairs of the 15,745 protein-coding genes. Next, we applied modularity maximization using the Louvain community detection algorithm^22^ and a consensus clustering algorithm^23^ to the 15,745 × 15,745 gene co-expression network. We identified four gene modules with distinct spatial distributions of gene expression levels (**Fig. 4A**): 1) a frontotemporal-dominant module (#1) involving medial and lateral frontotemporal regions; 2) a cingulo-sensory-dominant module (#2) involving the middle and posterior cingulate cortices; 3) a posterior occipitoparietal-dominant module (#3) involving the visual cortex, parietal regions, and somatosensory cortex; 4) a medial frontoparietal-dominant module (#4) involving the anterior and isthmus cingulate cortices, medial prefrontal cortex, and lateral frontal regions. In both cohorts, module 4 showed consistent negative associations between the average gene expression profile and the regional vulnerability to Aβ deposition (**Fig. 4B**). In the IMAS cohort, module 2 also showed significant negative gene-to-Aβ associations. In both cohorts, module 1 and 2 showed consistent positive and negative gene-to-tau associations, respectively. In the IMAS cohort, module 3 also showed a significant negative gene-to-tau association.

**Figure 4.**
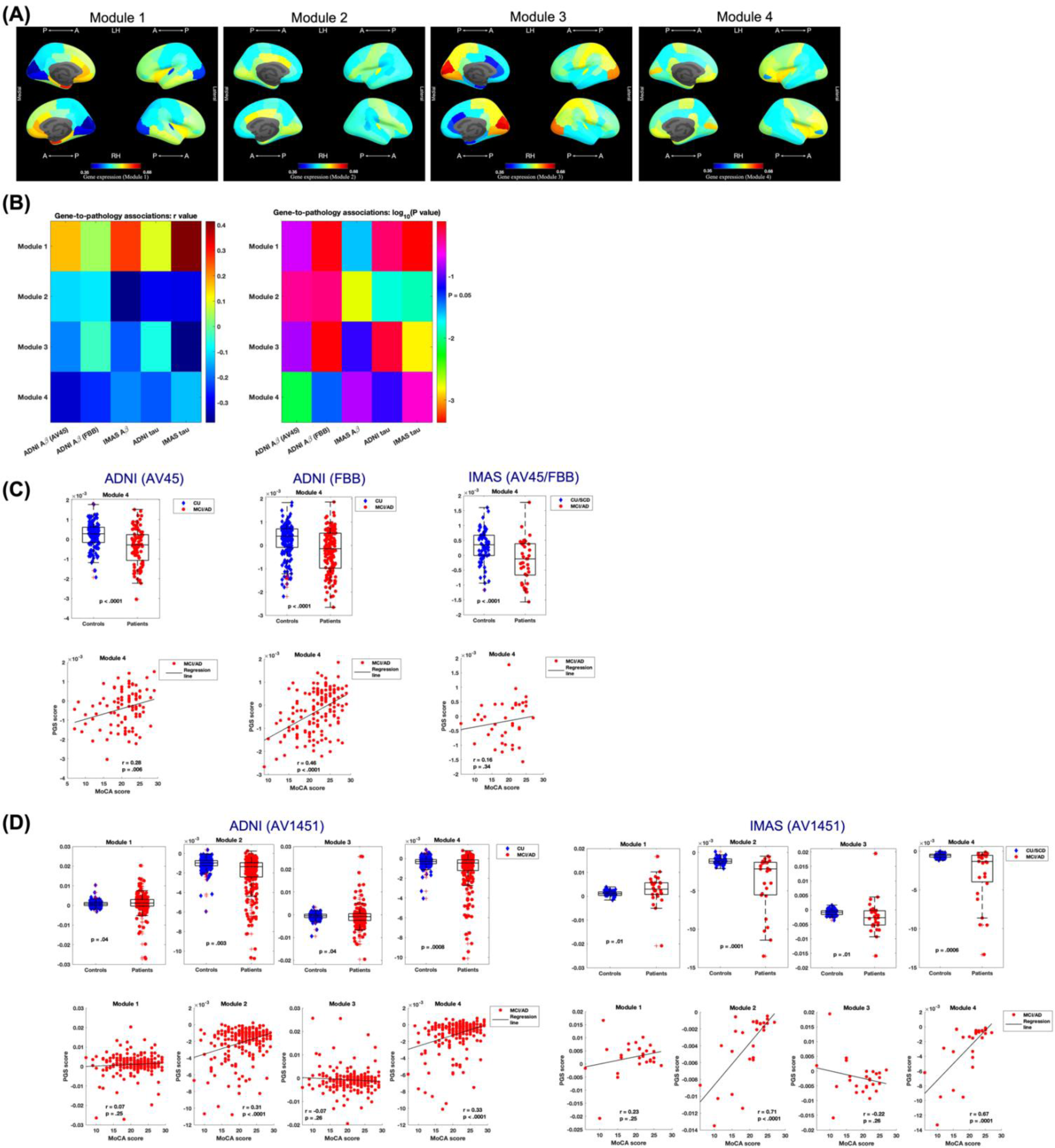
Four gene modules identified in the gene co-expression network showing distinct spatial patterns **(A)**. Spatial gene-to-Aβ and gene-to-tau associations **(B)** between average regional gene expression profiles of the four gene modules and regional vulnerability to Aβ and tau pathologies across the two cohorts: ADNI (AV45), ADNI (FBB), IMAS (AV45/FBB), ADNI tau (1AV451), and IMAS tau (1AV451). *P*-values were log10 transformed in **(B)**. Group differences of pathogenetic scores (PGSs) between patients and controls and their relationships to cognitive dysfunction in AD for the Aβ **(C)** and tau **(D)** pathologies, respectively. Abbreviations: AD = Alzheimer’s disease; IMAS = Indiana Memory and Aging Study; ADNI = Alzheimer’s Disease Neuroimaging Initiative; AV1451 = [^18^F]flortaucipir; AV45 = ^18^F]florbetapir; FBB = [^18^F]florbetaben.

### Biochemical pathways related to gene-to-pathology associations

In a secondary data-driven analysis, we computed gene-to-Aβ and gene-to-tau associations for all the 15,745 genes. We then used explorative GSEA to identify potential biochemical pathways of gene sets (genes annotated by the same gene ontology (GO) term including biological processes and cellular component functions) related to the gene-to-pathology associations. We identified 4 negatively enriched gene sets (e.g., peptide antigen processing and transmembrane protein complex) associated with the gene-to-Aβ associations and 11 positively enriched gene sets (e.g., cytosolic ribosome, synaptic and postsynaptic functions, and axoneme assembly) related to the gene-to-tau associations that were consistently shown in both cohorts. **Fig. 5** and Fig. **6** show representative gene enrichment plots for negatively and positively enriched GO gene sets with highest normalized enrichment scores for the gene-to-Aβ and the gene-to-tau associations, respectively. A detailed list of negatively and positively enriched gene sets identified by the GSEA is provided in **Supplement Table S2**.

**Figure 5.**
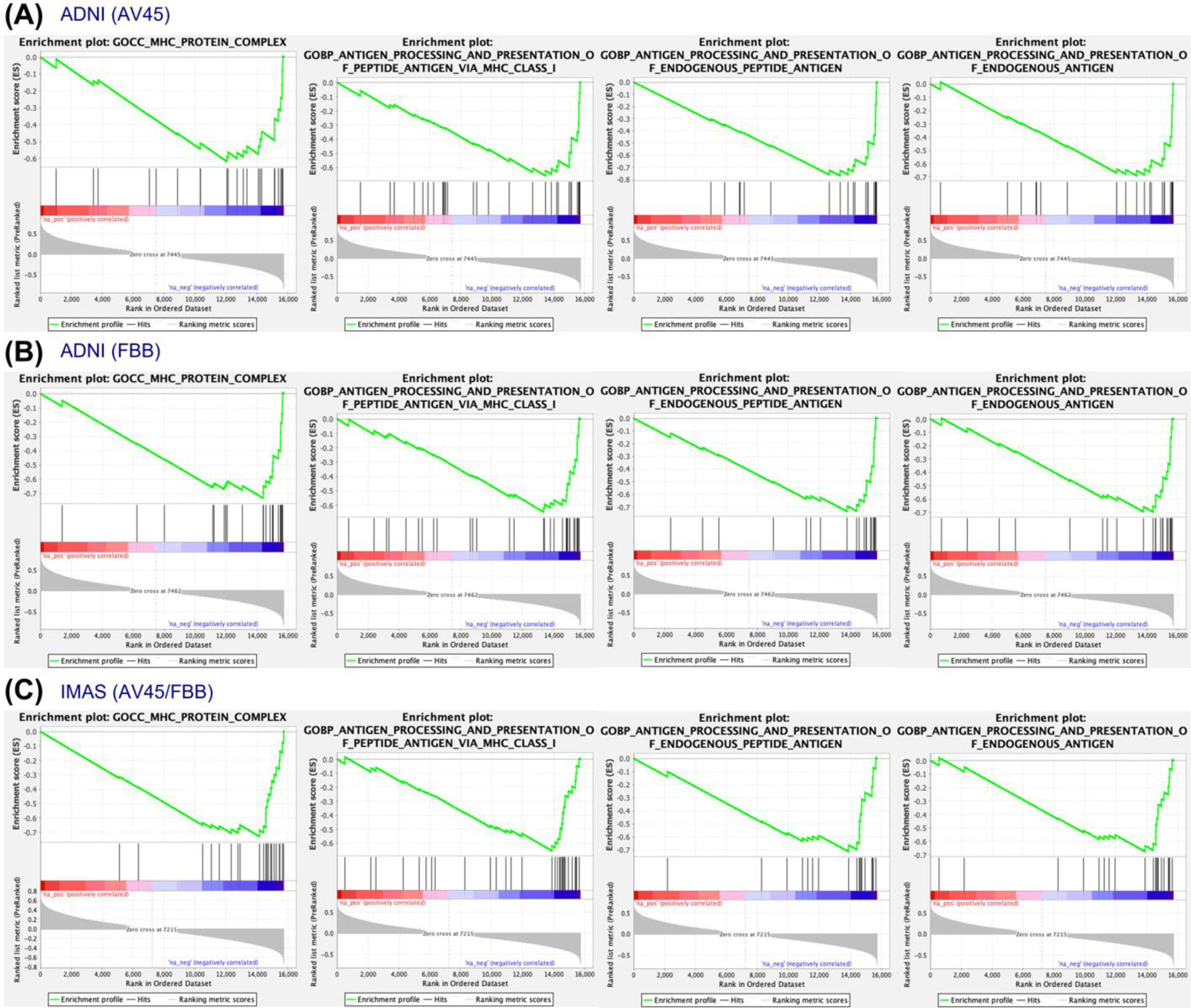
Enrichment plots for genes that were negatively associated with the gene-to-Aβ association across the two cohorts: ADNI (AV45) **(A)**, ADNI (FBB) **(B)**, and IMAS (AV45/FBB) **(C)**. Abbreviations: MAS = Indiana Memory and Aging Study; ADNI = Alzheimer’s Disease Neuroimaging Initiative; AV45 = [^18^F]florbetapir; FBB = [^18^F]florbetaben.

**Figure 6.**
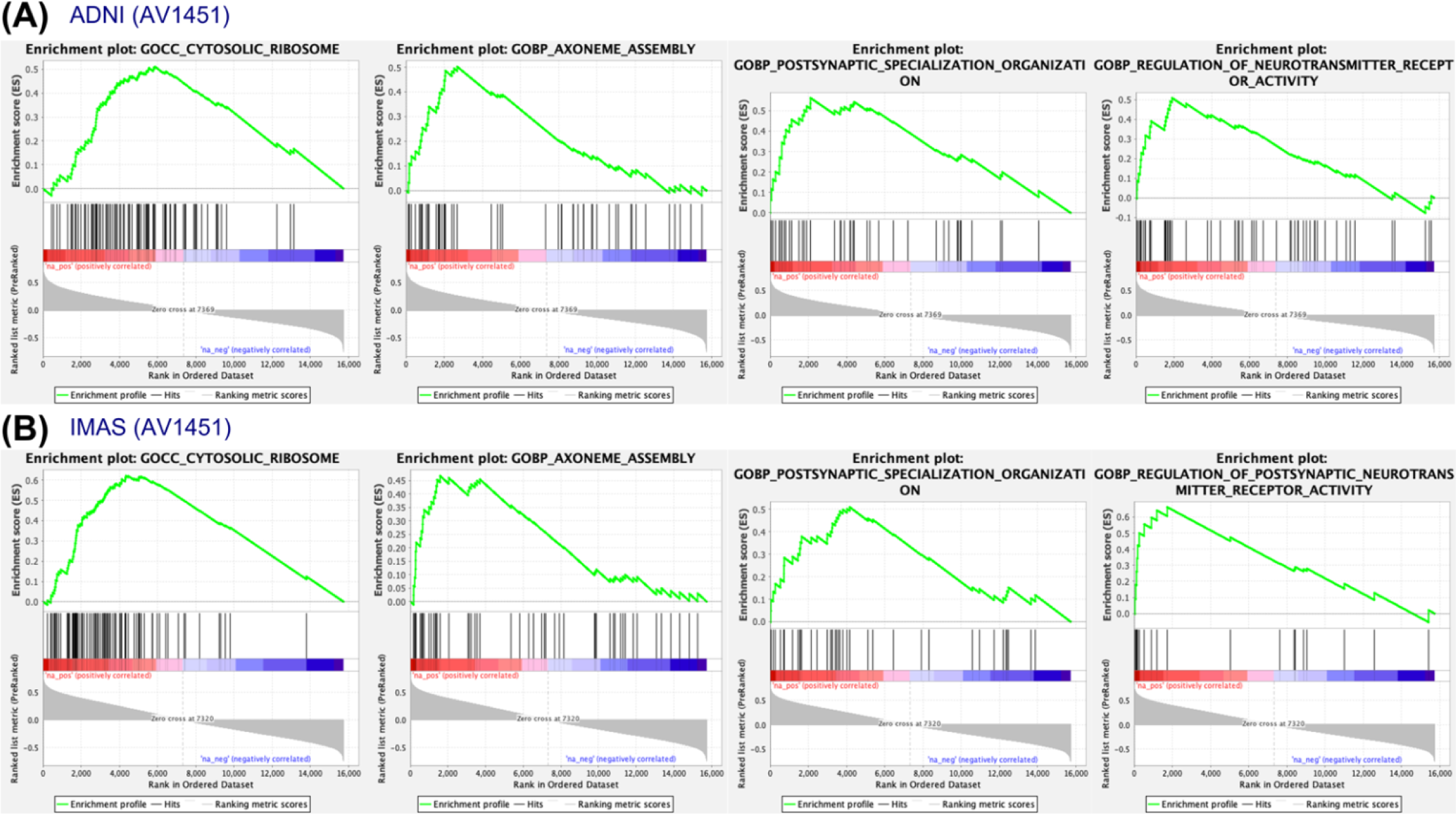
Enrichment plots for genes that were positively associated with the gene-to-tau association across the two cohorts: ADNI (AV1451) **(A)** and IMAS (AV1451) **(B)**. Abbreviations: IMAS = Indiana Memory and Aging Study; ADNI = Alzheimer’s Disease Neuroimaging Initiative; AV1451 = [^18^F]flortaucipir.

### Clinical implication of gene-to-pathology associations

We developed a novel analytic framework to study how the identified gene-to-pathology associations are related to cognitive decline in AD. Specifically, we first computed individual-level gene-to-pathology covariance values (named as pathogenetic scores [PGSs]) by estimating the covariance between regional gene expression profiles for each AD candidate gene (or gene module) and regional AD pathology (e.g., Aβ or tau deposition) values across all the individuals. Of note, this analysis was restricted to those AD candidate genes and gene modules that showed significant spatial correlations with the regional vulnerability to Aβ or tau pathologies (gene-to-Aβ and gene-to-tau associations) described above. Then, for those genes (or gene modules) showing significantly different PGCSs between patients (MCI and AD) and controls (CU and/or SCD) (*P*_FDR_ < 0.05), we estimated Pearson’s correlations between individual-level PGSs and MoCA performance. The PGSs and their correlations to the MOCA values were calculated for Aβ (e.g., the ADNI AV45 and FBB data and IMAS Aβ data) and for tau (e.g., ADNI and IMAS tau data) data included in this study, respectively.

For both cohorts, the PGSs showed consistent group differences (patients vs. controls) for Aβ-related (**Fig. 7**) and tau-related (**Fig. 8**) individual genes and gene modules (**Fig. 4C, D**). For Aβ-related genes (i.e., *CNTNAP2* and *HLA-DRB1*) and gene module (e.g., module 4), the PGSs showed comparable ranges of distributions between patients and controls, whereas for tau-related genes (i.e., *MAPT* and *APOE*) and gene modules (e.g., modules 2 and 4), the PGSs showed wider distributions in patients relative to controls. For both Aβ-related and tau-related genes and gene modules, the PGSs with significantly higher values in patients than in controls were negatively associated with MoCA total score in patients (i.e., for Aβ-related genes, see *CNTNAP2* in **Fig. 7 A, B, C**; for tau-related genes, see *MAPT* in **Fig. 8A, B**). Alternatively, the PGSs with significantly lower values in patients than in controls were positively correlated with MoCA total score in patients (i.e., for Aβ-related genes, see *HLA-DRB1*in **Fig. 7A, B, C**; for tau-related genes, see *ADAMTS4* in **Fig. 8A, B**; for Aβ- and tau-related gene modules, see **Fig. 4C, D**). Similar relationships between the gene-to-pathology covariance (PGSs) and cognitive decline were observed using the MMSE instead of the MoCA in the ADNI cohort (see **Supplement Table S3**).

**Figure 7.**
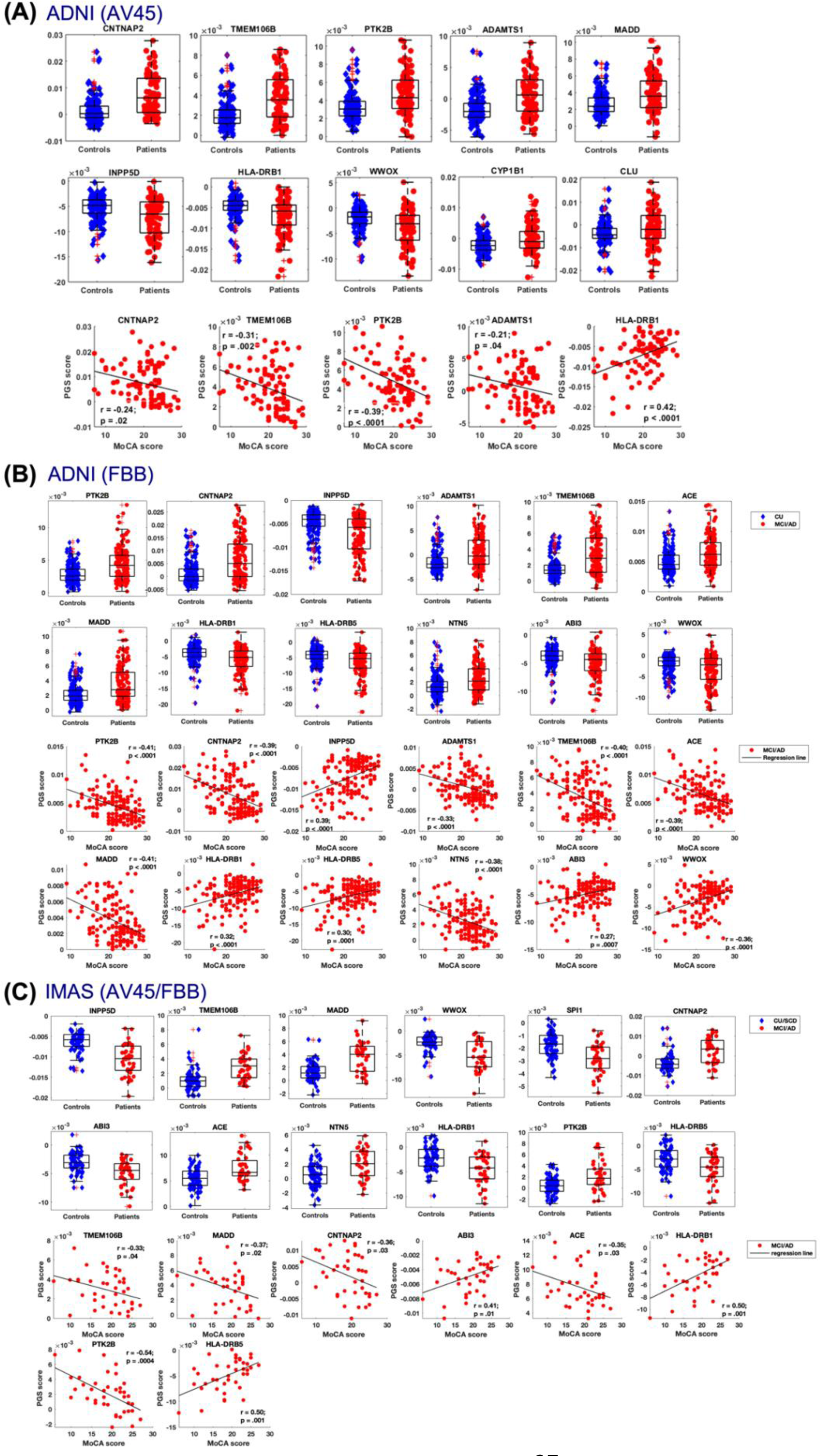
Group differences of Aβ-related pathogenetic scores (PGSs) between patients and controls and their relationships to cognitive dysfunction in AD across the two cohorts: ADNI (AV45) **(A)**, ADNI (FBB) **(B)**, and IMAS (AV45/FBB) **(C)**. Abbreviations: CU = cognitively unimpaired; SCD = subjective cognitive decline; MCI = mild cognitive impairment; AD = Alzheimer’s disease; IMAS = Indiana Memory and Aging Study; ADNI = Alzheimer’s Disease Neuroimaging Initiative; AV45 = [18F]florbetapir; FBB = [18F]florbetaben.

**Figure 8.**
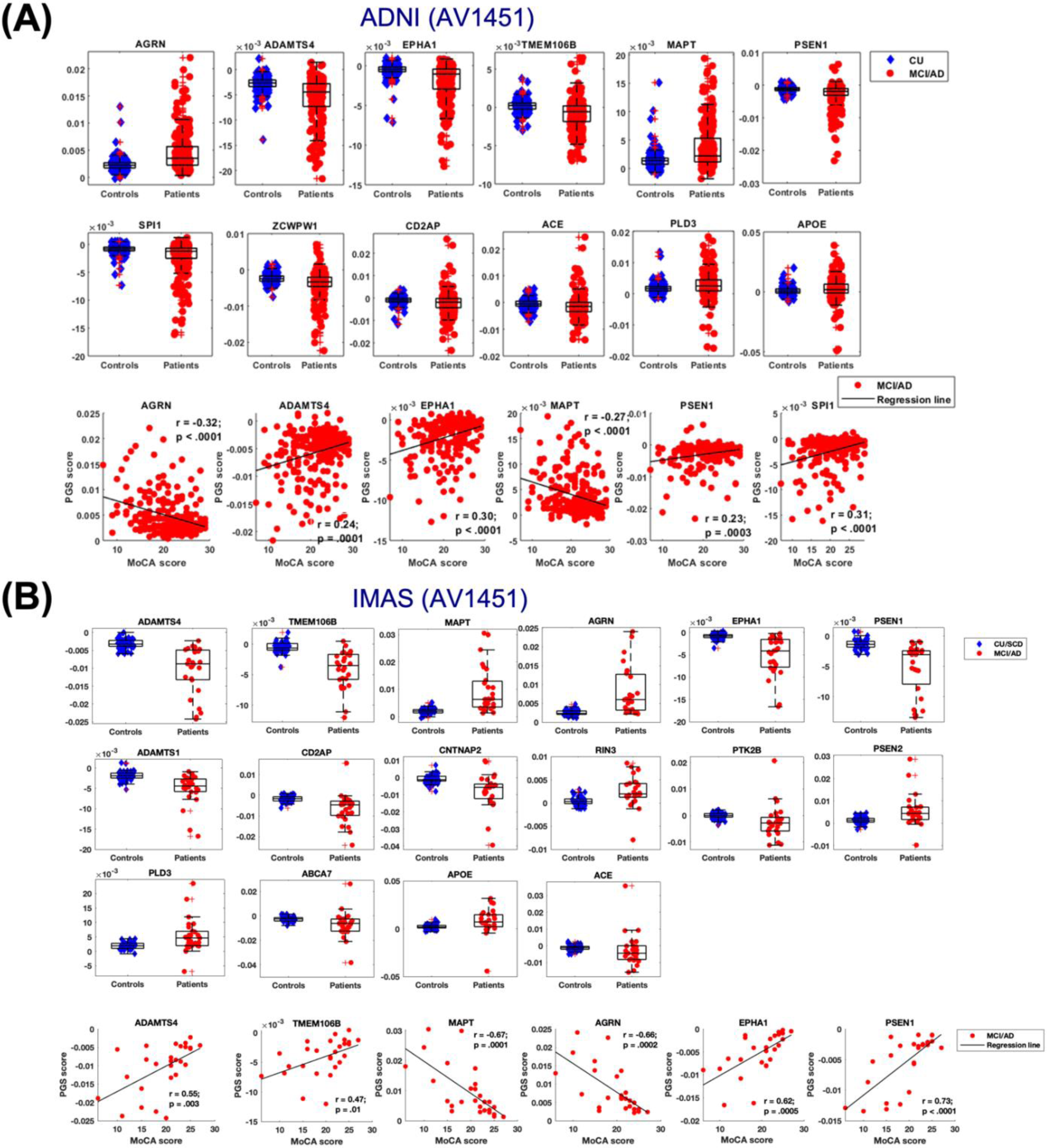
Group differences of tau-related pathogenetic scores (PGSs) between patients and controls and their relationships to cognitive dysfunction in AD across the two cohorts: ADNI (AV1451) **(A)** and IMAS (AV1451) **(B)**. Abbreviations: CU = cognitively unimpaired; SCD = subjective cognitive decline; MCI = mild cognitive impairment; AD = Alzheimer’s disease; IMAS = Indiana Memory and Aging Study; ADNI = Alzheimer’s Disease Neuroimaging Initiative; AV1451 = [^18^F]flortaucipir.

## DISCUSSION

In this study, we confirmed the presence of distinctive spatial vulnerability patterns of Aβ and tau pathologies and identified their relationships to regional gene expression profiles of specific AD susceptibility genes and gene co-expression network modules. In addition, the identified gene-to-Aβ and gene-to-tau associations were related to distinct biochemical pathways. Moreover, we identified relationships between the gene-to-pathology (e.g., Aβ and tau) covariance and level of cognitive impairment in AD, which supports the clinical implication of understanding these Aβ- and tau-related gene-to-pathology associations. Of note, these findings were validated and successfully replicated using PET imaging data (for Aβ, AV45 vs. FBB; for tau, AV-1451) and two cognitive measures (MoCA vs. MMSE) from two large independent cohorts.

Previous studies suggested the significant roles of spatial *MAPT*^16^ and *APOE*^17,24^ expression profiles in facilitating brain-wide tau spreading in healthy aging. In line with these findings, we found that gene expression patterns of both *MAPT* and *APOE* were related to regional vulnerability to tau deposition in patients with MCI and AD compared to CU individuals. Of note, in addition to these consistent findings, our study also found that the positive *APOE*-to-tau association was specific to AD, whereas the positive *MAPT*-to-tau association was shared in both AD and FTLD patients, suggesting that there might be common genetic basis underlying the regional vulnerability of tauopathy. *APOE* has been frequently associated with Aβ pathology^25–27^, but seldom with tau pathology in human studies^28^ (but see a mouse model study^29^). Our study highlights the significant role of regional *APOE* expression pattern in relation to regional tau pathology. One recent study^30^ found that *APOE*2 was related to lower regional tau burden, whereas *APOE*4 was related to higher regional tau deposit in Aβ positive CU, suggesting divergent effects of *APOE*2 and *APOE*4 on regional tau pathology in preclinical AD. The possible different spatial effects of *APOE* alleles (*APOE*2, *APOE*3, and *APOE*4) to tau pathology need to be considered in future studies.

In addition to *MAPT* and *APOE*, we identified novel positive and negative gene-to-tau associations for recently discovered AD risk genes from large GWAS studies. For instance, our finding of an AD-specific negative *CD2AP* gene-to-tau association agrees with the findings from a recent study^31^, where the CD2AP protein (a CD2-associated scaffold protein) was found to be colocalized with phosphorylated tau in AD brains through immunofluorescence analysis. This study also found a strong and positive association between CD2AP immunodetection in neurons and Braak neurofibrillary tangle stage. Moreover, previous studies^32,33^ have reported a link between *CD2AP* expression and tau pathology as measured by cerebrospinal fluid (CSF) tau biomarkers. Finally, among all the tauopathy-related genes detected, the *ADAMTS4* showed the strongest sex-independent gene-to-tau association in both AD and FTLD. It has been suggested that *ADAMTS4*, mainly expressed in oligodendrocytes, can produce strongly aggregating Aβ forms^34^, but only a few studies have found a direct relationship between the *ADAMTS4* expression and tau pathology. Emerging research suggests that *ADAMTS4* expression may have an indirect effect on tau production^35^. For instance, *ADAMTS4* exhibits isoform-specific cleavage of Reelin, and the absence of Reelin has been linked to elevated tau phosphorylation and widespread formation of neurofibrillary tangles (NFTs^36,37^). Finally, *AGRN* and *PLD3* showed strong positive gene-to-tau associations in AD. However, the potential biological mechanisms underlying these brain-wide spatial associations remain unclear. The *AGRN* gene encodes the AGRIN protein, which plays important role in the regulation of axonal and dendritic growth^38^. One study^38^ found that AGRIN upregulated the expression of tau and other microtubule-associated proteins in hippocampal neurons, suggesting a potential link between *AGRN* expression and tau pathology. The *PLD3* gene encodes a lysosomal protein that is highly enriched in axonal spheroids in the hippocampus and cortex^39^. Of note, a recent study^40^ observed that neuronal overexpression of *PLD3* affected axonal spheroids and induced neural circuit abnormalities in AD, while the deletion of *PLD3* reduced endolysosomal vesicle accumulation and improved neural network function. Accumulating evidence^1,41,42^ suggests that tau protein spreads via neural circuits, the processing of which might be related to the neuronal overexpression of *PLD3*^40^. Although these identified tauopathy-related genes and corresponding gene-to-tau associations are promising to advance our understanding of AD pathophysiology, future human and animal studies are required to validate our findings.

Findings of gene-to-Aβ associations are less convergent than those of gene-to-tau associations. One cross-sectional study^15^ found an *APP*-related gene-to-Aβ association, while another longitudinal study^16^ reported a *CLU*-related gene-to-Aβ association. Our study found gene-to-Aβ associations with *CLU* and *APP*, as well as other newly discovered Aβ-related genes whose regional expression patterns are related to the regional vulnerability of Aβ pathology. In particular, *CNTNAP2* and *TMEM106B* showed stronger positive gene-to-Aβ associations than *CLU*, *APP*, and other genes. Our findings are supported by previous studies. For example, one study^43^ found that *CNTNAP2* (a major gene in autism manifestation^44,45^) expression was downregulated in the hippocampus of AD patients. Moreover, three recent studies^46–48^ reported that amyloid fibrils in human brains, the main component of amyloid plaques, are formed by TMEM106B (a lysosomal/endosomal membrane protein), supporting our finding regarding the relationship between elevated *TMEM106B* expression and amyloid pathology. Of note, in addition to positive gene-to-Aβ associations identified by previous studies, we also found negative gene-to-Aβ associations, particularly involving regional *INPP5D* expression. Our previous studies^49,50^ have reported that *INPP5D* expression was increased in late-onset AD and showed a positive correlation with amyloid plaque density. Preclinical studies in mice with *INPP5D* deficiency was sufficient to regulate microglial functions and reduce Aβ pathology^50^. However, the previous study with clinical samples was completed in multiple brain regions with only select regions having increased *INPP5D* expression^49^, suggesting that some regions may be more vulnerable than others. Furthermore, in the previous study, we separated MCI from AD samples, which may also have additional implications on the findings. Therefore, additional studies in mice may be needed to determine which areas of the brain have altered *INPP5D* expression and if that relates to amyloid deposition.

We observed sex-dependent differences in regional vulnerability to Aβ and tau pathologies, which are in line with recent findings^51–56^. Interestingly, these sex-dependent regional differences were only seen in negative gene-to-Aβ and gene-to-tau associations, but not positive associations. The potential biological mechanisms that might drive sex-dependent gene-to-pathology associations is unclear, although changes in endogenous or exogenous estrogen levels during menopausal stage in females might provide a possible explanation^52^. For instance, studies have linked Aβ^57–59^ and tau^52,60^ pathologies with changes in estrogen levels. Moreover, animal studies^61^ have also found a direct relationship between tau vulnerability and estrogen depletion post-menopause. An alternative explanation may come from the sex differences in dysregulated immune responses^62^. Supporting this hypothesis, one study^63^ found that microglial nuclei isolated from aged female donors exhibited an increased expression of AD risk genes, gene signatures associated with the inflammatory response in AD, and genes linked to proinflammatory immune responses, compared to microglial nuclei from male donors. Taken together, these findings indicate that the susceptibility of specific brain regions to Aβ and tau pathology may be influenced by both the sex-independent overexpression and sex-dependent under-expression of multiple genes and their associated regulatory processes.

Due to the polygenic nature of sporadic late-onset AD, accumulation of Aβ and tau pathologies is unlikely driven by regional expression patterns of individual AD risk genes. To test this hypothesis, we evaluated the joint contribution of multiple genes through two data-driven analyses using the whole AHBA gene data. Of note, we found that the discordance between the localization of Aβ and tau pathologies in the brain could be explained by average gene expression profiles of distinct gene co-expression network modules. On the one hand, the spatial vulnerability of Aβ pathology was related to a medial frontoparietal-dominant gene expression pattern. Notably, the medial frontoparietal regions are parts of the default mode network (DMN^5,64^) derived from resting-state functional MRI data, which often show abnormal neural activity and connectivity in patients with AD^4,6,65,66^.

Converging evidence indicates that Aβ initially accumulates in the medial frontoparietal DMN regions^1,67^. Therefore, our findings might provide a genetic basis for the colocalization of abnormal Aβ accumulation and abnormal DMN connectivity in AD. On the other hand, we found that spatial vulnerability of tau pathology was related to the regional expression patterns of three gene modules. It should be noted that a previous study^68^ identified four distinct subtypes of tau accumulation patterns, some of which share similar spatial patterns with the expression patterns of the gene modules identified in the present study. Thus, one explanation could be that individuals with a specific tau accumulation subtype might be determined by the regional expression pattern of a specific gene module subtype. For instance, individuals with limbic-predominant tau pattern might have a cingulo-sensory-dominant gene expression profile, whereas individuals with posterior tau pattern might have a posterior occipitoparietal-dominant gene expression profile. These findings support our hypothesis that the regional average expression patterns of multiple genes (not only the individual AD susceptibility genes) might play vital roles in shaping the regional vulnerability patterns of Aβ and tau pathologies. Future studies are needed to test if the four subtypes of tau accumulation patterns are specifically related to the regional expression patterns of the four gene modules we identified in this study.

The GSEA results suggest that the positive and/or negative gene-to-Aβ and gene-to-tau associations at the levels of individual genes and gene modules are related to distinct biochemical pathways. Specifically, in line with previous studies, we found that the regional vulnerability to Aβ pathology was related to low expression levels of gene sets implicated in peptide antigen processing and transmembrane protein complex, whereas the regional vulnerability to tau pathology was associated with high expression levels of genes implicated in cytosolic ribosome, synaptic and postsynaptic functions, and axoneme assembly. The distinct biochemical pathways underlying the regional vulnerability to Aβ and tau pathologies might also explain the discordance between the localization of Aβ and tau pathologies in the brain^15,69–73^.

Since the development of the AHBA, a growing body of evidence suggests that specific regional gene expression profiles are related to brain structure and function in healthy subjects^13,74–90^ and neuropathology-related abnormalities of brain structure and function in multiple neurological and neuropsychiatric disorders, such as frontotemporal dementia^91^, Parkinson’s disease^92–96^, schizophrenia^97–100^, blind children^101^, early-life trauma^102^, major depression^83,103,104^, and autism^105^. However, these gene-to-connectivity or gene-to-pathology associations have not been translated to clinical utility. To fill this gap, the present study develops an innovative step forward by assessing the clinical significance of the identified gene-to-pathology associations in AD. Specifically, we developed an analytic framework characterizing individualized gene-to-pathology covariance that was related to cognitive dysfunction in patients with AD. Of note, not only can this framework aid in the translation of pathogenetic associations relevant to AD into clinical applications, but it can also be extended to understand cognitive function in healthy subjects and cognitive dysfunction in other brain disorders as mentioned above.

Some limitations of this study should be noted. First, our discoveries are primarily derived from correlation analyses, and as such, do not establish a causal relationship between gene expression patterns and the selective vulnerability of specific brain regions to Aβ and tau pathologies. Future studies using longitudinal designs may help identify possible causal relationships of these pathogenetic processes. Animal models could also be used to verify our findings. Second, the brain-wide gene expression data were measured postmortem in brains of six CU adults (aged 24–57 years; 5 males/1female) from the AHBA. To our knowledge, the AHBA is the only source of gene expression data currently available for brain-wide spatial association analysis. Future studies using age- and sex-matched individuals with CU aging or AD patients are required to replicate our findings. Last, the current study focuses on the detection of transcriptomic factors that contribute to the regional vulnerability of Aβ and tau pathologies, which cannot explain how Aβ and tau interact. Both human^106,107^ and transgenic mice^108,109^ studies have suggested that Aβ could remotely facilitate tau spreading from the MTL to other brain regions (e.g., the precuneus), possibly through long-range structural connections, leading to presumed downstream consequences, such as neurodegeneration and cognitive decline. However, it remains unknown if specific gene expression profiles might be playing a significant role in these pathological processes and if these processes might differ at different disease stages. Future studies leveraging longitudinal multimodal imaging data, including brain connectivity, regional gene expression profiles, as well as Aβ and tau PET data, may be promising to resolve these important questions.

Overall, these results support the hypotheses that specific regional gene expression profiles are related to the selective regional vulnerability of deposition of Aβ and tau pathologies in AD, and that distinct biochemical pathways are involved in the gene-to-Aβ and gene-to-tau associations. Taken together, they support the corollary hypothesis^1,110,111^ that gene transcription-associated regional vulnerability could explain the spatial discrepancies of Aβ and tau accumulation patterns that are observed in AD. Moreover, our findings highlight the clinical implication of the individualized gene-to-pathology associations in patients with AD, bridging the gap between pathogenetic mechanisms and clinical implication. The spatial gene-to-pathology associations and their relationships to cognitive decline in AD were replicated across two large independent datasets with different PET tracers and clinical measures. In the future, these findings and others may allow for forecasting and monitoring disease progression in individual patients, and ultimately help identify novel individualized pathogenetic biomarkers and therapeutic targets.

## MATERIALS AND METHODS

### Study design

The main aim of this study is to identify relationships between regional gene expression profiles and regional vulnerability to Aβ and tau pathologies in AD. Leveraging two large AD datasets, the ADNI and the IMAS cohorts, we tested three hypotheses: 1) regional expression profiles of distinct individual genes and gene modules are associated with regional vulnerability to Aβ (gene-to-Aβ associations) and tau (gene-to-tau associations) pathologies; 2) distinct biochemical pathways are related to specific gene-to-Aβ and gene-to-tau associations; 3) the identified gene-to-Aβ and gene-to-tau associations are linked to cognitive dysfunction in individual AD patients. For both cohorts, participants were included based on availability of Aβ and tau PET and a structural T1-weighted MRI. All eligible participants from both observational studies were included in the analysis. Sample sizes were not predetermined in advance, and due to the nature of both cohorts, randomization of participants was not possible. In the ADNI cohort (http://adni.loni.usc.edu/), we included 583 participants (328 CU, 192 MCI, 25 AD) with Aβ PET data and 564 participants (312 CU, 190 MCI, 62 AD) with tau PET data. In the IMAS cohort, we included 110 participants (38 CU, 31 SCD, 30 MCI, 11AD) with Aβ PET data and 78 participants (30 CU, 20 SCD, 23 MCI, 5 AD) with tau PET data. We also included 11 patients with frontotemporal lobar degeneration (FTLD; 6 with behavioral variant FTD [bvFTD] and 5 with primary progressive aphasia [PPA]) from the Indiana Alzheimer’s Disease Research Center (IADRC) cohort. We computed gene-to-tau associations for these FTLD patients and tested if the identified gene-to-tau associations were specific to AD or shared with FTLD. A multidisciplinary consensus meeting was held to make diagnoses. To be eligible for participation, individuals must not have an active psychiatric or neurological disorder. The specific inclusion/exclusion criteria for the ADNI cohort can be found at http://www.adni-info.org. The Indiana University Institutional Review Board approved the study, and written informed consent was obtained from each participant from both the IMAS and ADNI cohorts according to the Declaration of Helsinki.

### Neurocognitive variables

All participants underwent a comprehensive clinical assessment and neuropsychological battery, as described in previous ADNI^112,113^ and IMAS^114,115^ studies. In this study, we used total scores of the Montreal Cognitive Assessment (MoCA) and the Mini-Mental State Exam (MMSE) for characterizing the cognitive function of participants and for the purpose of replicating results. Of note, the MoCA and the MMSE were used in the ADNI cohort, while only the MoCA was used in the IMAS cohort.

### Image acquisition and preprocessing

Participants from both cohorts had an anatomical MRI with whole brain coverage using a 3D Magnetization Prepared Rapid Gradient Echo (MPRAGE) sequence (220 sagittal slices, 1.1 × 1.1 × 1.2mm^3^ voxels) per the Alzheimer’s Disease Neuroimaging Initiative (ADNI-2) imaging protocol. In the IMAS cohort, we implemented an accelerated protocol (GRAPPA, R=2) to reduce imaging time from 9:14 s (IMAS dataset) to 5:12 s. All the T1-weighted images were preprocessed using FreeSurfer v6.0 (https://surfer.nmr.mgh.harvard.edu/) as previously described^116–118^.

### Aβ and tau PET imaging preprocessing

All participants from the IMAS and ADNI had two PET imaging acquisitions: (1) [^18^F]flortaucipir (18F-AV-1451) PET scan, a tracer that binds to tau in neurofibrillary tangles and neurites; and, (2) one of two amyloid PET tracers, [^18^F]florbetapir (AV45) or [^18^F]florbetaben (FBB) that bind to fibrillary Aβ plaques. The ADNI Aβ and tau PET data were processed by Dr. William Jagust’s lab^119–122^ and were downloaded from the publicly available database at the Laboratory of Neuro Imaging (LONI) Image & Data Archive (IDA) Repository (https://ida.loni.usc.edu/). Briefly, each Aβ PET scan was co-registered to the MRI closest in time. A native-space MRI scan for each subject is processed (segmentation and parcellation) with FreeSurfer version 6 to define a cortical summary region. The cortical summary region consists of frontal, anterior/posterior cingulate, lateral parietal, and lateral temporal regions. Summary SUVRs were calculated by normalizing the cortical summary region by the FreeSurfer-defined whole cerebellum (a reference region) with a positivity threshold of 1.11 for the [^18^F]florbetapir SUVRs and a positivity threshold of 1.08 for the [^18^F]florbetaben SUVRs. Similar processing procedures were applied when generating summary Flortaucipir SUVRs with Braak stage composite regions as the cortical summary region and inferior cerebellar gray matter as the reference region^123,124^. Partial volume correction (PVC) was not applied to the [^18^F]flortaucipir SUVRs, as previous studies have rarely found PVC-related effects in gene-to-tau associations^16,17^.

The details of IMAS PET data processing analyses were described in previous studies^116,125,126^. PET data were reconstructed using an ordered subset expectation maximization algorithm with weighted attenuation following Siemens manufacturers protocols. Using SPM12, all PET data in native space were corrected for motion, co-registered to their corresponding T1-weighted MRI images, and spatially normalized into MNI152 template using normalization parameters obtained from the T1-weighted MRI normalization. Static images from 50-70 minutes or 90-110 minutes were created as the sum of appropriate time frames for [^18^F]florbetapir or [^18^F]florbetaben, respectively. Static images were intensity normalized to the whole cerebellum to create standardized uptake value ratio (SUVR) images. For [^18^F]flortaucipir, static images were created from 80-100 minutes post-injection and intensity normalized to the cerebellar crus to create SUVR images. Finally, all PET scans were smoothed with an 8mm full width half maximum Gaussian kernel. Median Aβ and tau PET SUVR values were extracted from FreeSurfer’s Desikan-Killiany (D-K) atlas defined 68 cortical regions^127^. Aβ positivity was defined as a SUVR > 1.1 for [^18^F]florbetapir and SUVR > 1.2 for [^18^F]florbetaben. Continuous variables for PET images were used in the subsequent association analysis.

### Regional transcriptional analysis

Regional gene expression profiles for 20,736 protein-coding genes were derived from brain-wide microarray-based transcriptome data from the Allen Human Brain Atlas^11,12^. The microarray probes were collected from 3,700 regional brain tissue samples in autopsy data of six adult individuals (5 males/1 female; aged 24–57 years) without history of neurological disorders. Of note, as the first two donors (1 male and 1 female) did not show interhemispheric asymmetries and sex-related differences in gene expression data, the subsequent four donors (all males) had brain tissue collection in the left hemisphere only. A recommended analysis pipeline^14,128^ was used to preprocess the gene expression data in the left hemisphere, including probe-to-gene re-annotation, data filtering, probe selection, sample assignment, gene filtering, and normalization across the 6 donors. Specifically, the following probe filtering criteria were applied: i) the probe-to-gene annotations were updated using Re-Annotator package; ii) the reannotated probes with expression measures lower than the background in more than 50% samples were discarded; iii) a representative probe with the highest intensity was selected to represent a gene. This procedure retained 15,745 probes, each representing a unique protein-coding gene. Gene expression values were normalized separately for each donor across cortical regions and then averaged across donors. The D-K atlas was used to parcellate each gene expression map into 34 cortical regions in the left hemisphere. Of note, we used the D-K atlas to define the same cortical regions for both the PET and gene expression imaging data. Based on the hemispheric symmetry of gene expression patterns, the left hemisphere regional gene expression values for each gene were mirrored to the right hemisphere^17^, resulting in gene expression values of 68 cortical regions for each gene. Finally, a gene expression matrix (15,745 × 68) was constructed, where rows of the matrix correspond to the 15,745 genes and columns correspond to 68 cortical regions.

### Hypothesis-driven analysis

#### Candidate gene analysis

We tested the mechanistic hypothesis that the regional vulnerability to Aβ and tau pathologies are spatially correlated with regional expression levels of AD candidate genes associated with Aβ and tau pathology. As a first step, we estimated the regional vulnerability to Aβ and tau pathologies by contrasting Aβ and tau PET imaging data between cognitively normal participants (CU in the ADNI cohort; CU and SCD in the IMAS cohort) and patients (MCI and AD in two cohorts) using independent samples *T*-test. The similarities of regional vulnerability to Aβ and tau pathologies between the ADNI and IMAS cohorts were estimated by computing Pearson’s correlations between the *T*-statistic scores of the two cohorts. Then, we computed spatial associations between regional gene expression levels and regional vulnerability to Aβ (gene-to-Aβ associations) and tau (gene-to-tau associations) pathologies (represented as case-control *T*-statistic maps), for 60 AD susceptibility genes selected from recent large-scale GWAS studies^18–20^. Following preprocessing, 15 genes were excluded due to not meeting the quality control criteria outlined above, resulting in 45 out of the initial 60 genes being eligible for candidate gene analysis. The Benjamini-Hochberg’s False Discovery Rate (B-H FDR) correction (*P*_FDR_ < 0.05) was used to control for false positive results caused by multiple comparisons.

#### Spatial permutation test (spin test)

Recent studies^129^ have found that statistical significance of spatial correlations between imaging maps could be inflated by failing to consider the spatial autocorrelation effects: neighboring data points are unlikely statistically independent. In this study, we use Vasa’s spin test method^130^ to control for the inherent spatial autocorrelation effects in the PET imaging and transcriptomic data. Briefly, a spatial permutation framework was used to generate null spatial models by randomly spinning (or rotating; number of rotations = 10,000) the spherical representations of the parcellated cortical map (e.g., D-K atlas) and preserving the spatial relationships across brain regions^131^. The reconstructed gene expression data were used to generate a null distribution of correlation coefficients, which were then used to evaluate if the observed spatial correlations exceed the expected null spatial correlations estimated by using randomized brain regions with the same neighboring relationships.

#### Gene specificity test

We assessed the gene specificity by comparing the observed spatial gene-to-Aβ and gene-to-tau associations to a null distribution of associations estimated by other sets of genes^132^. Specifically, we constructed a null model to test if the observed associations will be stronger than the null distribution of associations estimated by randomly selected background genes that are significantly overexpressed in the brain tissues than in other body sites.

#### Pathology specificity analysis

We assessed if the regional vulnerability to tau pathology and its relation to regional gene expression profiles were specific to AD or shared with frontotemporal lobar degeneration (FTLD), a different dementia type sometimes characterized by tauopathy^133–137^. Specifically, we included 11 patients with FTLD (6 with behavioral variant FTD [bvFTD] and 5 with primary progressive aphasia [PPA]) from the IADRC cohort and added 11 FTLD risk genes^138–144^ (e.g., *C9orf72*, *GRN*, *FUS*, *KIAA0319*, *VCP*, *TARDBP*, *CHMP2B*, *ITM2B*, *TBK1*, *TBP*, *CTSF*) to the analysis. We then reconducted the regional tau PET analysis and regional transcriptional analysis as described above.

#### Sex specificity analysis

Previous studies reported sex-dependent differences in regional vulnerability to Aβ^58,59^ and tau^51–53,55,60^ pathologies. To assess the potential effects of sex on the identified spatial gene-to-Aβ and gene-to-tau associations, we recomputed the regional vulnerability to tau pathology and its relation to regional gene expression profiles in males and females, separately. Due to sample size issues, we focus on the ADNI Aβ and tau data when conducting the sex specificity analysis.

### Data-driven analysis

#### Gene co-expression network analysis

Genes often show correlated spatial expression pattens, indicating that they do not function independently^11,12,77,145^. Thus, going beyond hypothesis-driven analysis using a prior set of selected individual AD candidate genes, we performed a data-driven analysis linking the regional vulnerability to Aβ and tau pathologies with average expression profiles of gene co-expression modules consisting of hundreds or thousands of genes. Specifically, we first constructed a weighted gene co-expression network (15,745 × 15,745) by computing Pearson’s correlations between the regional expression profiles for pairs of the 15,745 genes. In this gene co-expression network, each gene is a node and each link between two nodes is the Pearson correlation between the regional expression profiles of each pair of genes. The correlation coefficients were then Fisher r-to-z transformed. Next, we used the Louvain’s community detection method^22^ (gamma = 1.5; symmetric treatment of negative weights) and a consensus clustering algorithm^23^ (τ = 0.4; repetition times = 50) implemented in the Brain Connectivity Toolbox^146,147^ to identify consensus gene co-expression modules (or clusters). Of note, we included links with both positive and negative values into the community detection analysis, as previous gene co-expression studies^148–150^ demonstrated the existence of negatively correlated gene expression, suggesting the equal importance of both positive and negative link weights in gene co-expression network. Subsequently, we estimated the average gene expression level for each gene co-expression module, and then computed gene-to-pathology associations between regional gene expression levels for each module and the regional vulnerability to Aβ and tau pathologies, respectively.

#### Gene set enrichment analysis

In a secondary data-driven analysis, we performed gene set enrichment analysis (GSEA^151,152^) to identify biochemical pathways of gene sets with expression patterns associated with the regional vulnerability to Aβ and tau pathologies. We used gene sets involved in biological processes and cellular component functions that group genes using annotations from Gene Ontology (GO^153^). First, we calculated spatial correlations between genome-wide regional expression profiles of all the 15,745 genes and the regional vulnerability to Aβ and tau pathologies, respectively. Next, we ranked the 15,745 genes according to their spatial correlation values: the top (positive correlations) and bottom parts (negative correlations) of this ranked list contain the genes of interest, expression values of which increase or decrease, respectively, in relation to the regional vulnerability to Aβ or tau pathologies. Then, GSEAPreranked analysis was performed against the ranked genes using the GSEA software (version 4.3.2). The normalized enrichment score was computed to quantify the non-random distribution of a gene set in a ranked list, while also taking into consideration the varying sizes of the functional gene sets being analyzed. Permutation testing (1000 permutations) was used to assess statistical significance. FDR correction (*P*_FDR_ < 0.05) was used to control for the independent testing of multiple gene sets. Finally, we performed a leading-edge analysis to analyze commonalities among the most relevant genes of the identified pathways by clustering the respective leading-edge gene subsets, the principal genes that account for a gene set’s enrichment signal.

### Clinical implication of gene-to-pathology associations

We developed a novel analytic framework for assessing the clinical implication of the identified gene-to-Aβ and gene-to-tau associations. Specifically, for the ADNI [^18^F]florbetapir data, we first computed covariance values between regional gene expression values for each AD candidate gene and individualized regional Aβ deposition values across all the individuals, resulting in individual-level gene-to-Aβ covariance values, named as Aβ-related pathogenetic scores (PGSs). Then, we estimated group differences of the Aβ-related PGSs between patients (MCI and AD) and cognitively normal controls (CU and/or SCD). Next, for the genes showing significantly different Aβ-related PGSs between groups (*P*_FDR_ < 0.05), we computed Pearson’s correlations between the PGSs and MoCA total score. This analytic framework links the gene-to-pathology (Aβ or tau) associations with cognitive dysfunction in AD, characterizing the clinical implication of gene-to-pathology associations. To assess their reproducibility, the PGSs and their correlations to MoCA performance were calculated for other Aβ (e.g., the ADNI FBB and IMAS Aβ data) and for tau (e.g., ADNI and IMAS tau data) data. The PGSs and their correlations to MoCA performance were calculated for both AD candidate genes and gene modules identified by gene co-expression network analysis. As the ADNI cohort also has MMSE data, we validated the relationships between the gene-to-pathology (e.g., Aβ and tau) covariance (PGCSs) and cognitive decline measured by MMSE total score in the ADNI AD patients.

### Statistical analysis

Statistical analyses of subject characteristics were performed with jamovi (version: 2.3.21; https://www.jamovi.org/download.html). Group differences in age, MoCA, and MMSE were tested using one-way ANOVA tests, while sex differences between groups were tested using a chi-square (χ^2^) test. Other statistical analyses were described detail above.

## SUPPLEMENTARY MATERIALS

**Supplement Table S1**. Gene-to-Aβ and gene-to-tau associations accounting for the effects of spatial autocorrelation and gene specificity.

**Supplement Table S2**. Negatively and positively enriched gene sets identified by the GSEA.

**Supplement Table S3**. Relationships between the gene-to-pathology covariance and cognitive dysfunction measured by MoCA and MMSE, respectively.

**Supplement Figure S1.** Spatial gene-to-tau associations between brain-wide gene expression profiles of AD/FTD susceptibility genes and brain-wide tau PET data in the IMAS cohort.

**Supplement Figure S2.** Spatial gene-to-Aβ associations between brain-wide gene expression profiles and brain-wide Aβ data measured in the males and females, separately.

**Supplement Figure S3.** Spatial gene-to-tau associations between brain-wide gene expression profiles and brain-wide tau data measured in the males and females, separately.

## Data Availability

The ADNI data are publicly available at Laboratory of Neuro Imaging (LONI) Image & Data Archive (IDA) Repository on the ADNI website. All data produced in the present study are available upon reasonable request to the authors.

## Acknowledgments

**Funding:** This work was supported by several U.S. National Institute of Aging (NIA) and Alzheimer’s Association (AA) grants: M.Y. is supported by grants from the Alzheimer’s Association (AARF-22-722571) and the National Institute on Aging (U19AG074879, R01 AG019771, P30 AG072976, U01 AG072177, and U01 AG068057).

## Author contributions

Conceptualization: M.Y., O.S., A.J.S Methodology: M.Y., O.S., S.R. Investigation: M.Y., S.R. Visualization: M.Y.

Funding acquisition: M.Y., A.J.S., K.N. Project administration: A.J.S., M.Y. Supervision: M.Y., A.J.S., O.S., S.R.

Writing – original draft: M.Y., O.S., A.J.S., S.R. Writing – review & editing: All the authors

## Data and materials availability

All the data associated with this study are present either in the paper or in the Supplementary Materials. ADNI data are available online after signing a data use agreement. The IMAS data management is overseen by A.J.S. and S.R. at the Indiana Alzheimer’s Disease Research Center (IADRC).

