## Supplemental Figures for "Spatial transcriptomic patterns underlying regional vulnerability to amyloid-β and tau pathologies and their relationships to cognitive dysfunction in Alzheimer’s disease"

**Supplement Figures**

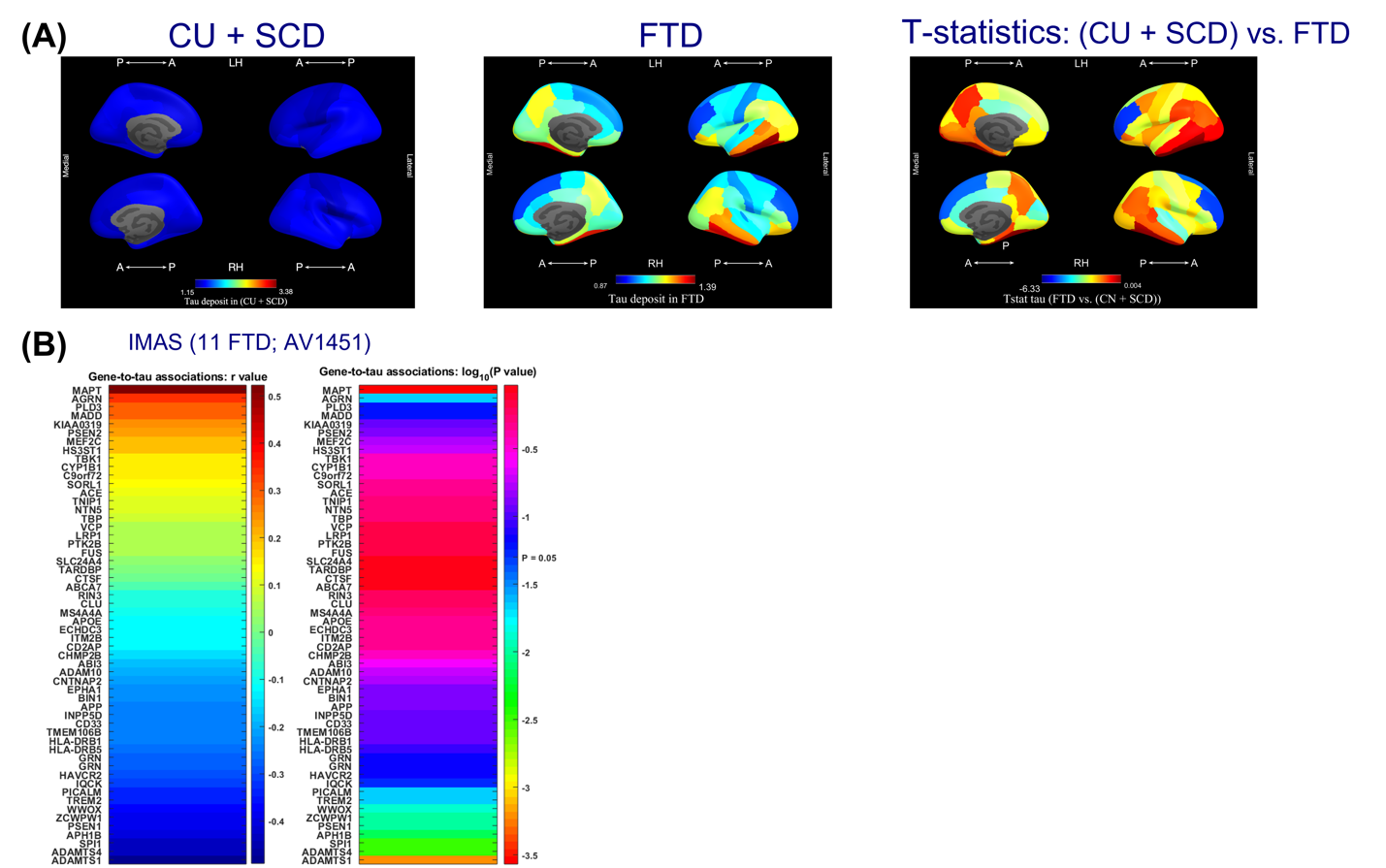

**Figure S1.** **(A)** Spatial patterns for tau deposits (SUVR values) across diagnosis groups and *T*-statistics map for regional differences. **(B)** Spatial gene-to-tau associations between brain-wide gene expression profiles of AD/FTD susceptibility genes and brain-wide tau PET data in the IMAS cohort. *P*-values were log10 transformed in **(B)**. Abbreviations: A = anterior; P = posterior; RH = right hemisphere; LH = left hemisphere; CU = cognitively unimpaired; SCD = subjective cognitive decline; IMAS = Indiana Memory and Aging Study; AV1451 = [^18^F]flortaucipir.

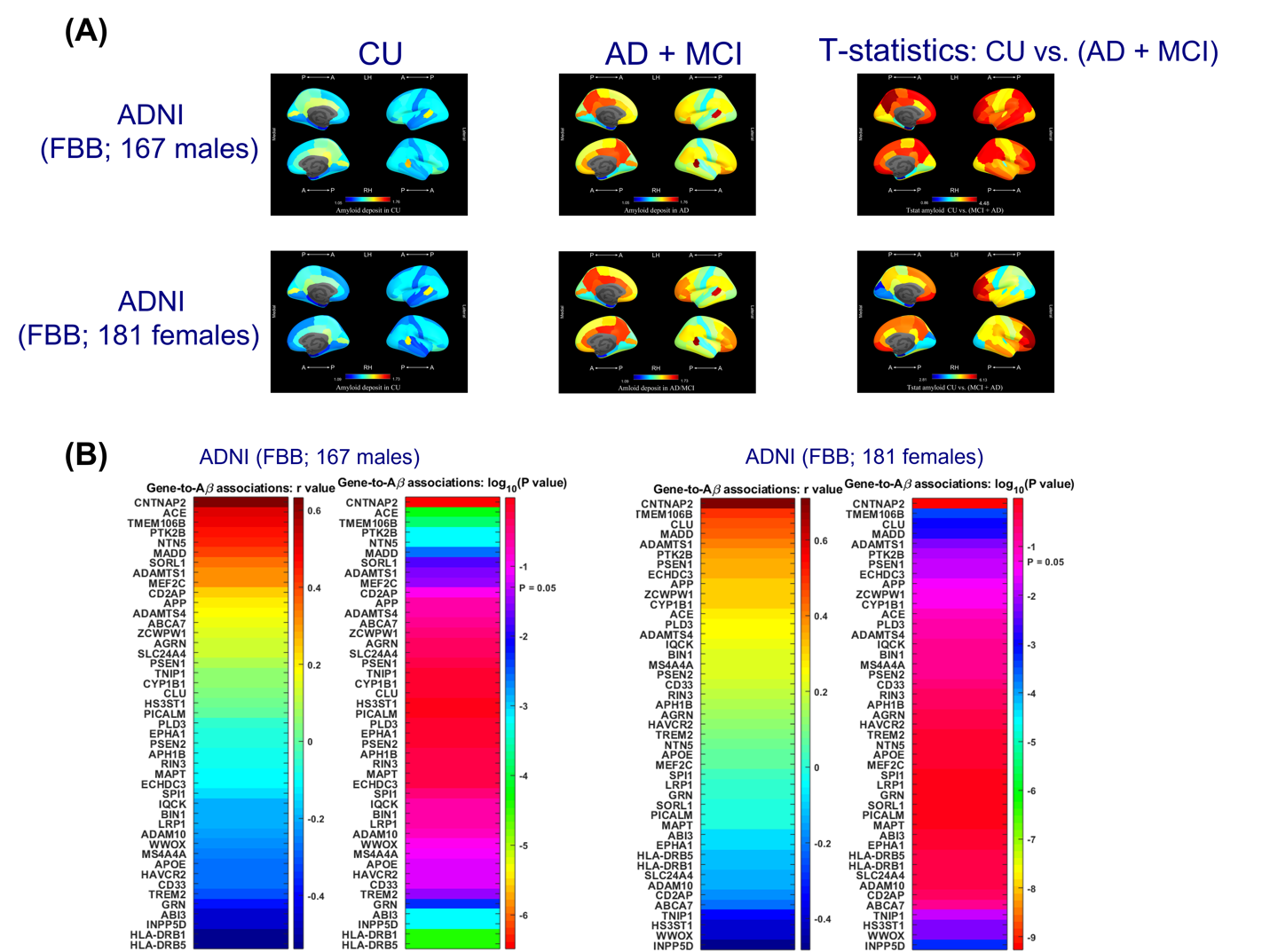

**Figure S2. (A)** Spatial patterns for Aβ deposits (SUVR values) across diagnosis groups and *T*-statistics map for regional differences in males and females, respectively. **(B)** Spatial gene-to-Aβ associations between brain-wide gene expression profiles and brain-wide Aβ data measured in the males and females, separately. *P*-values were log10 transformed in **(B)**. Abbreviations: A = anterior; P = posterior; RH = right hemisphere; LH = left hemisphere; CU = cognitively unimpaired; SCD = subjective cognitive decline; MCI = mild cognitive impairment; AD = Alzheimer’s disease; ADNI = Alzheimer’s Disease Neuroimaging Initiative; FBB = [^18^F]florbetaben.

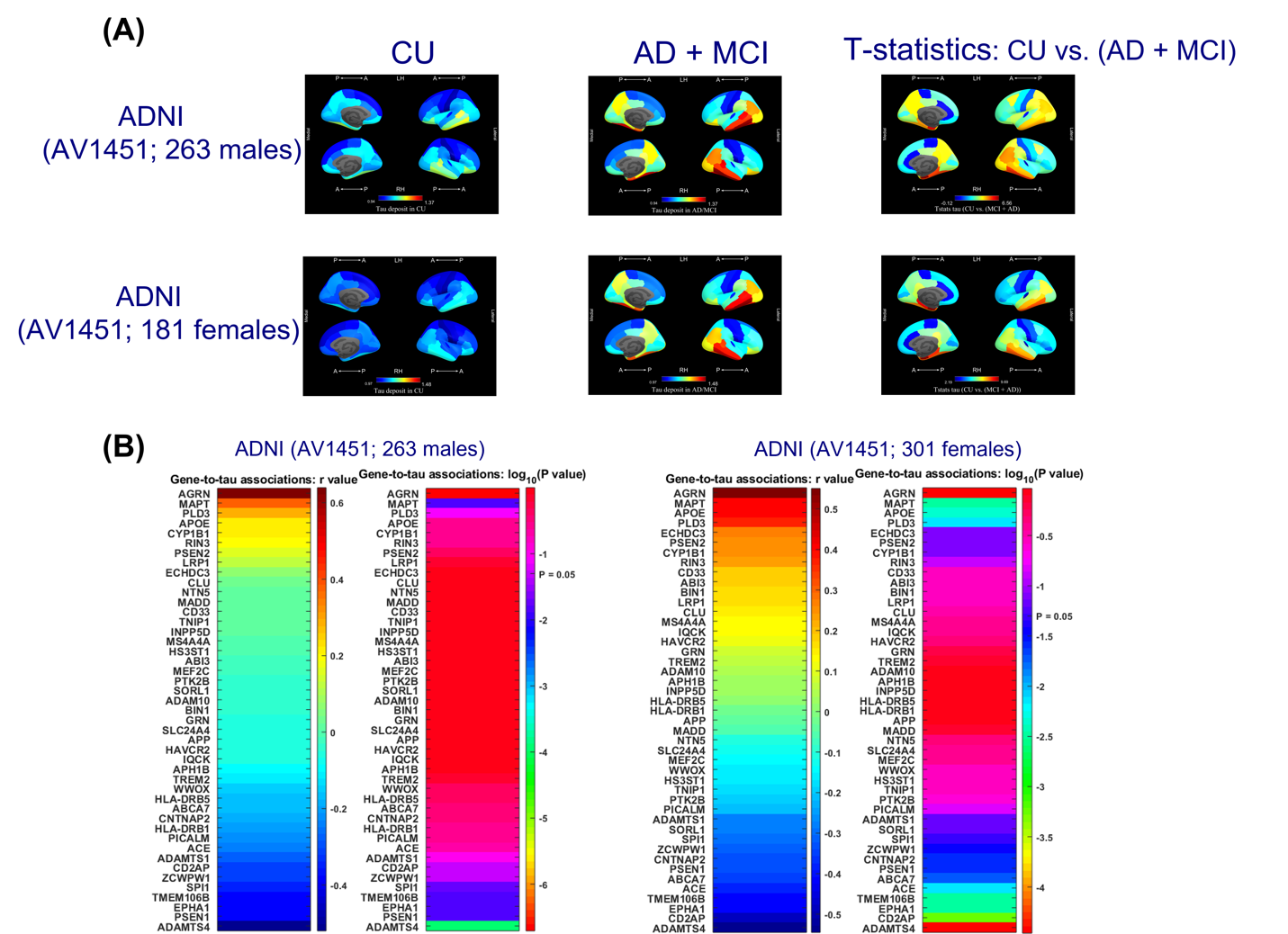

**Figure S3. (A)** Spatial patterns for tau deposits (SUVR values) across diagnosis groups and *T*-statistics map for regional differences in males and females, respectively. **(B)** Spatial gene-to-tau associations between brain-wide gene expression profiles and brain-wide tau data measured in the males and females, separately. *P*-values were log10 transformed in **(B)**. Abbreviations: A = anterior; P = posterior; RH = right hemisphere; LH = left hemisphere; CU = cognitively unimpaired; SCD = subjective cognitive decline; MCI = mild cognitive impairment; AD = Alzheimer’s disease; ADNI = Alzheimer’s Disease Neuroimaging Initiative; FBB = [^18^F]florbetaben.
